# Bronchoalveolar lavage metabolome dynamics reflect underlying disease and chronic lung allograft dysfunction

**DOI:** 10.1101/2022.11.16.22281980

**Authors:** Christian Martin, Kathleen S. Mahan, Talia D. Wiggen, Adam J. Gilbertsen, Marshall I. Hertz, Ryan C. Hunter, Robert A. Quinn

## Abstract

**Background:** Progression of chronic lung disease often leads to the requirement for a lung transplant (LTX). Despite improvements in short-term survival after LTX, chronic lung allograft dysfunction (CLAD) remains a critical challenge for long-term survival. This study investigates the relationship between the metabolome of bronchoalveolar lavage fluid (BALF) from subjects post-LTX with underlying lung disease and CLAD severity.

**Methods:** Untargeted LC-MS/MS metabolomics was performed on 960 BALF samples collected over 10 years from LTX recipients with alpha-1-antitrypsin disease (AATD, n=22), cystic fibrosis (CF, n=46), chronic obstructive pulmonary disease (COPD, n = 79) or pulmonary fibrosis (PF, n=47). Datasets were analyzed using machine learning and multivariate statistics for associations with underlying disease and final CLAD severity.

**Results:** BALF metabolomes varied by underlying disease state, with AATD LT recipients being particularly distinctive (PERMANOVA, *p*=0.001). We also found a significant association with the final CLAD severity score (PERMANOVA, *p*=0.001), especially those with underlying CF. Association with CLAD severity was driven by changes in phosphoethanolamine (PE) and phosphocholine lipids that increased and decreased, respectively, and metabolites from the bacterial pathogen *Pseudomonas aeruginosa. P. aeruginosa* siderophores, quorum-sensing quinolones, and phenazines were detected in BALF, and 4-hydroxy-2-heptylquinoline (HHQ) was predictive of the final CLAD stage in samples from CF patients (*R*=0.34; *p*≤0.01). Relationships between CLAD stage and *P. aeruginosa* metabolites were especially strong in those with CF, where 61% of subjects had at least one of these metabolites in their first BALF sample after transplant.

**Conclusions:** BALF metabolomes after LTX are distinctive based on the underlying disease and reflect final CLAD stage. In those with more severe outcomes, there is a lipid transition from PC to predominantly PE phospholipids. The association of *P. aeruginosa* metabolites with CLAD stages in LTX recipients with CF indicates this bacterium and its metabolites may be drivers of allograft dysfunction.

**Key messages:** Despite the high prevalence of CLAD among LTX recipients, its pathology is not well understood, and no single molecular indicator is known to predict disease onset. Our machine learning metabolomic-based approach allowed us to identify patterns associated with a shift in the lipid metabolism and bacterial metabolites predicting CLAD onset in CF. This study provides a better understanding about the progression of allograft dysfunction through the molecular transitions within the transplanted lung from the host and bacterial pathogens.

## Introduction

Lung transplantation (LTX) is a therapeutic option for patients who develop progressive and severe chronic lung disease [1]. However, chronic lung allograft dysfunction (CLAD) remains a major barrier to long-term LTX patient survival, as it affects 50% of patients at five years post-transplantation [2,3]. Bronchiolitis obliterans syndrome (BOS), a crucial phenotypic manifestation of allograft dysfunction [4,5], is used as a scaled measure of disease progression (i.e., CLAD stage) [6]. Notoriously a heterogenous disease, BOS severity scores were recently updated in 2019 [6] to better characterize the stages of severity experienced by LTX recipients. These CLAD stages can occur post-LTX in patients with different pre-LTX lung diseases such as alpha-1-antitrypsin deficiency disease (AATD), cystic fibrosis (CF), chronic obstructive pulmonary disease (COPD), and pulmonary fibrosis (PF) [7–10].

Bronchoalveolar lavage fluid (BALF) is a valuable sample type for studying pathological characteristics that develop after LTX due to the direct proximity of BALF sample origin to the site of stress and/or injury in the lung [11]. Metabolites detected within BALF can be indicative of physiological processes of disease progression, microbial lung burden, and possibly, allograft rejection [12,13]. However, there are few studies of BALF metabolomes describing the composition, microbial virulence factors, amino acids, lipids, and a myriad of pharmaceuticals [14–17].

Untargeted mass spectrometry (MS)-based metabolomics allows exploration of the chemical diversity associated with a wide range of biological samples [18,19]. However, navigating the diverse chemical data generated in untargeted metabolomics studies, in which a large proportion of detectable metabolites are unknown, remains challenging [18,19]. Advances in bioinformatic analyses of MS data have enabled a more comprehensive interpretation of biological information contained within these highly technical and large datasets. For example, the Global Natural Products Social molecular networking web platform (GNPS) has simplified the exploration of the chemical space of metabolomes. GNPS is a tandem mass spectrometry (MS/MS) automated data organizational tool that enables comparisons of MS/MS fragmentation patterns among samples to cluster and visualize related molecules in a spectral network [20,21]. GNPS can be paired with metabolite feature quantification algorithms to create a comprehensive workflow for untargeted metabolomics of clinical samples [20,22]. In this study, we applied this approach to 960 BALF samples collected longitudinally from 194 LTX recipients to determine whether post-transplant BALF metabolomes reflect the underlying disease diagnosis; and whether specific molecular signatures correlate with allograft dysfunction.

## Materials and Methods

### Study Design, Subjects, and BOS-Grading

Subjects with AATD, CF, COPD, and IPF who underwent lung transplantation at the University of Minnesota consented to have a portion of their post-LTX BALF used for research. BALF was collected over a 10-year period from 2002 to 2012 as a routine procedure per transplant protocol or if undergoing diagnostic BALF collection due to clinical indications, such as new radiographical changes, new respiratory symptoms, or a decrease in FEV_1_. Inclusion criteria were ≥ 18 years of age; diagnosis of AATD, CF, COPD, or PF; and receipt of a bilateral or single lung transplant. Exclusion criteria were < 18 years of age and the inability to provide consent and/or tolerate the BALF procedure. Clinical characteristics of subjects in this study, including disease progression scores and samples collected for each, are displayed in Table 1 and Table S1. BOS-grade was recorded for patients based on the changes in their forced expiratory volume in one second (FEV_1)_, initially determined using definitions of the International Society for Heart and Lung Transplantation (ISHLT) [23], and updated here to reflect current CLAD definitions [6]. The final CLAD stage was known for each patient, while some measures were determined after sample collection ceased. This protocol was approved by the University of Minnesota IRB (STUDY00004547).

**Table 1.** Clinical characteristics of subjects (total n=194) and samples (total n=960) collected post-LTX grouped by prior underlying disease – alpha-1 anti-trypsin deficiency (AATD), cystic fibrosis (CF), chronic obstructive pulmonary disease (COPD), and idiopathic pulmonary fibrosis (IPF). BALF sampling information, microbiology associated with *Pseudomonas aeruginosa*, age at transplant (LTX), sex ratio (female/male). The averages of the last recorded forced expiratory volume per 1 second expressed as a percent of predicted (%FEV_1_) and the CLAD stage are displayed based on pre-transplant diagnoses.

| **Clinical parameters** | **AATD** | **CF** | **COPF** | **IPF** |
| --- | --- | --- | --- | --- |
| *Number of subjects* | 22 | 46 | 79 | 47 |
| *Sex ratio (F/M)* | 0.69 (9/13) | 1.20 (25/21) | 1.20 (43/36) | 0.42 (14/33) |
| *Average of age at LTX*<br>*(range)* | 55 (42 – 55) | 36 (20 – 54) | 59 (44 – 74) | 58 (39 – 68) |
| *Average of last recorded*<br>*FEV<sub>1</sub> (range)* | 64 (18 - 96) | 69 (17 – 100) | 65 (23 – 100) | 68 (26 – 99) |
| *Average of last recorded*<br>*CLAD stage (0 to 4)* | 1.57 | 1.35 | 1.62 | 1.36 |
| *Number of subjects with *P.**<br>**aeruginosa* (%)* | 11 (50%) | 32 (70%) | 7 (5%) | 4 (9%) |
| *BALF samples* | 130 | 185 | 362 | 283 |
| *BALF average/subject*<br>*(range)* | 5.01 (2-12) | 4.02 (1-9) | 4.58 (2-9) | 6.02 (3-10) |
| *Average of sample collection*<br>*(years)* | 2.93 | 3.33 | 3.64 | 4.05 |

### Bronchoalveolar lavage sampling material

Bronchoalveolar lavage fluid (BALF) was collected via orotracheal or nasotracheal bronchoscopy. Sterile saline was instilled into a subsegmental bronchus after advancement and occlusion of the airway lumen. BALF was separated into ∼1.5 mL aliquots and frozen at -80 °C for further untargeted metabolomic analysis (see supplementary methods). All 960 samples were included for analysis of variation (ANOVA) among underlying disease types and associations with final CLAD stages.

### Organic extraction and LC-MS/MS analysis

Methanolic extracts of 960 BALF samples (50:50 v/v) were analyzed on a Thermo QExactive™ mass spectrometer coupled to a Vanquish Ultra-High-Performance Liquid Chromatography (UHPLC) system (ThermoFisher) (see supplementary methods) [21,24]. Subsequently, feature-based molecular networking (FBMN) was performed with a parent and fragment mass ion tolerance of 0.02 Da, a cosine score of 0.65, and a minimum matched peaks minimum of 4 [22]. FBMN is publicly available at https://gnps.ucsd.edu/ProteoSAFe/status.jsp?task=756456ac794d48b0bba80dbe28e0de66, and raw data files are available in the MASSIVE data repository as <u>MSV000085760</u>. Data curation consisted of removing metabolites detected in blanks and removal of known pharmaceuticals and their related nodes in the molecular network (see supplementary methods, Figure S1, and Table S2). Presence/absence frequency and rank abundance of all molecular features were calculated after creating a cutoff by summing the relative abundances of features above 10 x e^5^ abundance in samples obtained from subjects with AATD, CF, COPD, or IPF.

### Statistical analysis

Variations in metabolomic data associated with underlying disease type and the final CLAD stage for each subject were assessed using beta-diversity metrics and tested for significance with permutational multivariate analysis of variance (PERMANOVA). Metabolome variation was further assessed using random forest (RF) classification and regression analysis (numerical CLAD stages, 0 to 4). A Bray-Curtis dissimilarity matrix was calculated on the entire metabolome and used to generate principal coordinate analysis (PCoA) plots through the in-house tool ClusterApp and visualized in EMPeror [25]. PERMANOVA tests were performed for diseases and CLAD stages (as a categorical variable) with subject-source as an interacting factor to account for variation in the number of samples per subject. Post-hoc tests among disease types were performed with R packages Devtools and Vegan (pairwise-adonis) [26,27]. Variable importance plots from both RF approaches were used to identify metabolites driving the variation observed. Pearson correlations were used to determine relationships between metabolite abundance and final CLAD stages, as well as the correlation of *P. aeruginosa* metabolites with its relative abundance determined using 16S rRNA sequence data from the same samples. 16S rRNA sequence data were generated using bacterial genomic DNA extracted from each sample, sequencing the V4 region using Illumina MiSeq TruSeq 2×300 paired-end technology, and sequence analysis in R as previously described [28]. The relative abundance of *P. aeruginosa* was determined using DADA2 [29]. R packages random forest, vegan and ggplot2 were also used for these analyses [30–32] (see supplementary methods).

## Results

### Sample collection and clinical design

The two primary objectives for this study were: i) to determine if the metabolome of BALF collected after LTX was associated with the underlying pre-LTX lung diagnosis, and ii) if the data were associated with measures of CLAD severity across all subjects and within each underlying disease type. The dataset comprised longitudinal BALF samples (n = 960) collected over a 10-year period from subjects (n = 194) with one of four underlying conditions prior to transplant; AATD (n = 22), CF (n = 46), COPD (n = 79), and PF (n = 47). Clinical parameters and patient demographic information are presented in Table 1. Subjects developed CLAD at different times and to varying degrees during the collection period. The final BOS-grade of all subjects was known, even when this measure was recorded after BALF collection had ceased. Thus, metabolome data variation was tested against this final BOS-grade measure.

### BALF metabolome variation based on the underlying lung disease type pre-transplant

Since BALF samples were obtained via bronchoscopy, numerous drugs and xenobiotics were identified in the dataset. We reasoned that these molecules could confound underlying biologically significant trends, so they were removed from the metabolome data using Global Natural Product Social (GNPS) library searching and molecular networking to identify known and chemically related pharmaceuticals in the GNPS libraries. After filtering pharmaceuticals and sample contaminants (detected in controls), the entire BALF data set included 4755 molecular features, of which 361 had a spectral match to known compounds in GNPS libraries [21].

PCoA ordination of the entire dataset, colored by pre-transplant disease diagnosis, revealed significant differences between AATD, CF, COPD, and PF cohorts (PERMANOVA *F* = 3.91, *p* = 0.001, Figure 1a). Additionally, post-hoc testing showed significant pairwise differences between diseases (Table 2), suggesting that allograft metabolomes are dependent, in part, on the underlying disease of the transplant recipient. This unique disease signature was especially marked in subjects with AATD, as evidenced by its low random forest (RF) classification error (13.3%, Table S3) and separation by PCoA clustering (Figure 1a). We then used supervised RF classification analysis to identify metabolites that most strongly distinguished BALF samples by the underlying disease. The top 10 metabolites driving differences between groups included phenylalanine, phosphocholines, and other lipids. Notably, phenylalanine was particularly abundant in subjects with underlying AATD (pairwise p-value CF p = 2.9 e^-11^, COPD p = 4.7 e^-14^, and IPF p = 3.1 e^-13^) (Figure 1b, Figure S2).

**Table 2.** Post-hoc pairwise comparisons of BALF metabolome among different diseases (AATD, CF, COPD, and IPF).

| Pairwise Comparisons | F-value | p-value |
| --- | --- | --- |
| COPD and IPF | 26.5689 | 0.001 |
| COPD and CF | 2.1869 | 0.002 |
| COPD and IPF | 1.7698 | 0.017 |
| AATD and CF | 21.2079 | 0.001 |
| AATD and IPF | 28.6926 | 0.001 |
| CF and IPF | 3.1142 | 0.001 |

**Figure 1.**
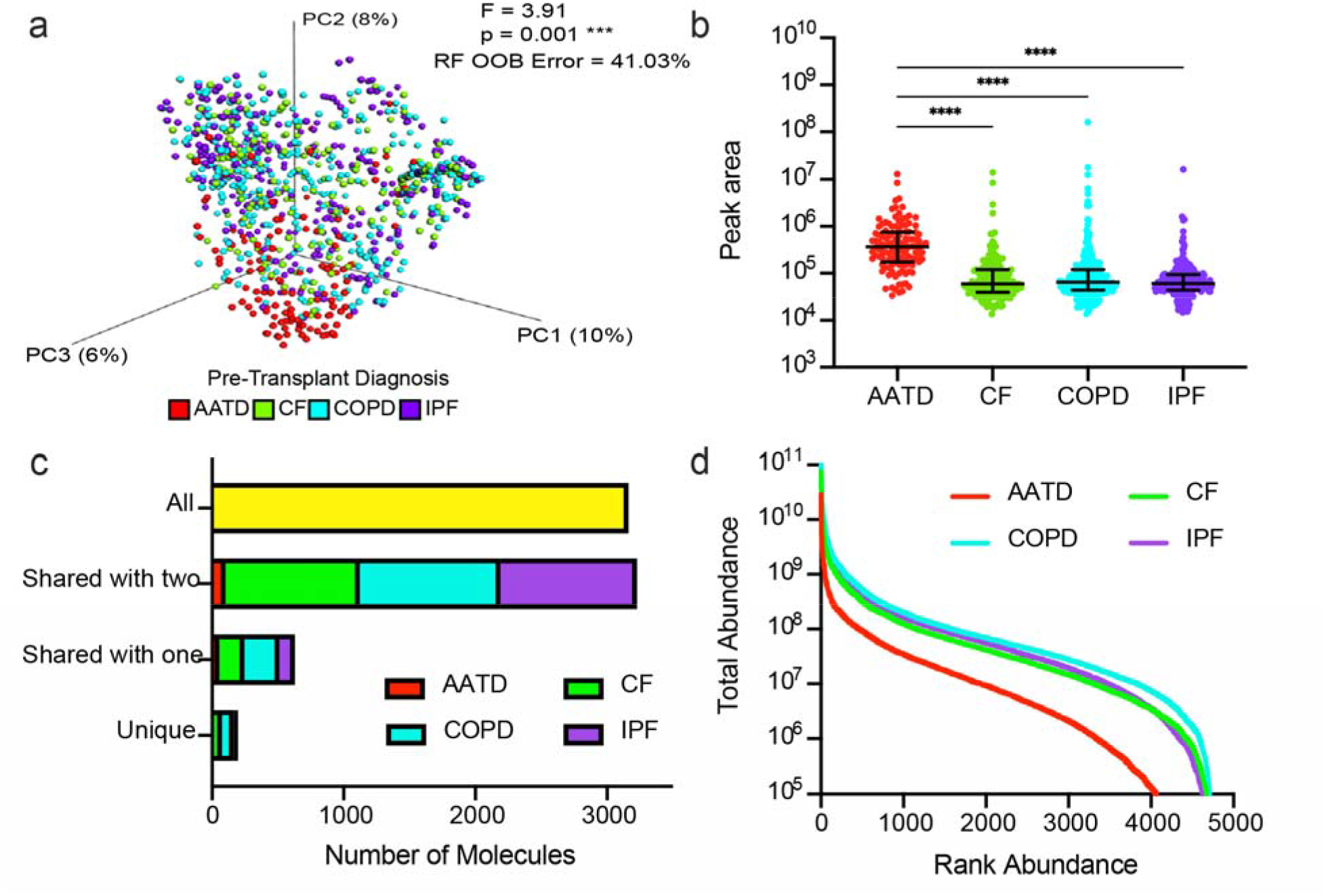
Lung allograft metabolomic profiles vary by pre-LTX condition. a) PCoA plot of a Bray-Curtis distance matrix of lung allograft BALF metabolomic profiles. Samples colored by pre-LTX disease and demonstrate significant differences among disease types (PERMANOVA, F=3.91, p = 0.001, random forest out-of-bag (OOB) error = 41.03%). b) Feature abundance of DL-Phenylalanine by disease state. Kruskal-Wallis test and post-hoc Dunn test p-values are shown. c) Uniqueness and sharing of BALF metabolites across diseases. Molecule presence or absence was determined and plotted by the number of molecules that are unique, shared with one other, two others, or all four diseases. d) Ranking molecular abundance curves of BALF metabolome colored by disease.

To further understand the nature of the unique BALF metabolome in LTX recipients with AATD, we analyzed metabolite presence/absence to determine the degree of chemical sharing among underlying diseases. Although most metabolites were shared across all four diseases, AATD unexpectedly had fewer molecules shared with the other diseases (either one other, or two others, Figure 1c). Rank abundance curves of the metabolome revealed that AATD had fewer overall metabolites, and these were less abundant than the other three disease types (Figure 1a,d).

### Longitudinal allograft metabolomes are associated with CLAD severity outcomes

Each subject’s final CLAD stage was then used to assess the relationship between the entire longitudinal metabolome dataset and disease severity (n = 960). PERMANOVA (categorical CLAD stage, F = 4.023, p = 0.001) and RF regression (linearized CLAD stage, variance explained = 11.08%) revealed an overall association of the final CLAD stage with collective longitudinal metabolome composition (Figure 2a). When tested separately on each pre-LTX disease diagnosis, CLAD stages maintained their categorical significance for all but IPF (Figure 2b). RF regression showed that LTX recipients with underlying CF had the strongest association of their BALF metabolome variation with numerical CLAD stages (% variance explained = 34.04), followed by COPD (14.36) and markedly less variation in IPF (4.65) and AATD (3.16) (Figure S3). These results demonstrate a significant relationship between the BALF metabolome after LTX and the final CLAD stage, which was particularly strong among individuals with CF.

**Figure 2.**
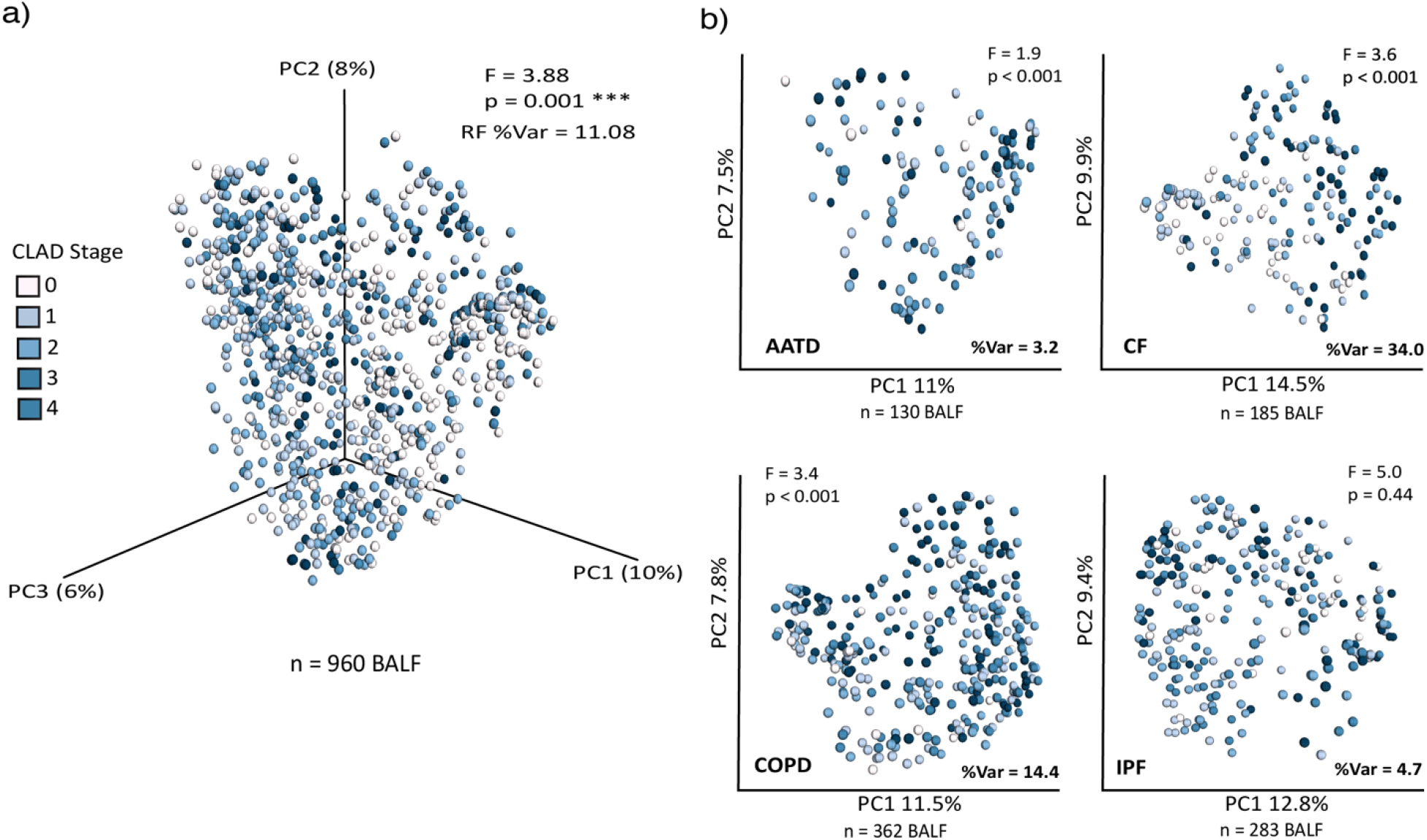
Principal coordinate analysis plots of Bray-Curtis distance matrices calculated for BALF metabolome based on CLAD stage measurements: Statistics from the categorical PERMANOVA testing (F and p-value) and linear variation based on the RF analysis (%Var) are shown for each plot. a) PCoA plot of the entire BALF metabolome colored by the final CLAD stages. b) PCoA plots of the entire BALF metabolomic data separated by disease type colored by the final CLAD stages.

### Altered phospholipids and *P. aeruginosa*-derived molecules drive association with the final CLAD stage

The association between CLAD stage and longitudinal metabolome composition, particularly among CF subjects, motivated further analyses of specific metabolites that most strongly influenced trends in the dataset. Variable importance plots from the RF analysis of the entire dataset and each disease analyzed separately revealed several metabolites of interest (Figure S3, S4). Molecular networking was then used to help annotate and identify these and related molecules. With this approach, we identified four clusters (‘molecular families’) of interest, including phosphoethanolamine (molecular family I), phosphocholine (molecular families II and III), and quinolone-like-molecules (molecular family IV, Figure S5). The molecular family I included the phospholipids lysophosphoethanolamine (lysoPE) 18:0/0:0 (*m*/*z* 482.3225 [M+H]^+^, C_23_H_49_NO_7_P, 3.4 ppm error) and lysoPE 18:1/0:0 (*m*/*z* 480.3073 [M+H]^+^, C_23_H_47_NO_7_P, 2.40 ppm error). Molecular family II included phosphatidylcholine 16:0/14:0 (*m*/*z* 728.5183 [M+Na]^+^, C_38_H_76_NO_8_P, 2.42 ppm error). Molecular family III corresponds to PC 16:0/18:0 (*m*/*z* 780.5529 [M+Na]^+^, C_42_H_80_NO_8_P -2.20 ppm error). Finally, in molecular family IV, we identified 4-hydroxy-2-heptylquinolone (HHQ, *m*/*z* 244.1689 [M+H]^+^, C_16_H_22_NO 2.8 ppm error), a *Pseudomonas aeruginosa-*derived quinolone and known quorum sensing metabolite that plays an integral role in the gene expression and physiology of this opportunistic pathogen (Figures S5 and S6) [33].

We then plotted the relationship between the feature abundances of each metabolite of interest with the final CLAD stages across the entire dataset. Both lysoPE(18:0/0:0) and lysoPE(18:1/0:0) significantly increased with disease progression (Pearson r = 0.23, *p* = 6.9e-13, and *r* = 0.16, *p* = 0.017, respectively). In contrast, phosphocholine molecules PC (16:0/14:0) and PC(16:0/18:0) decreased as final disease severity increased (Pearson *r* = -0.22, *p* = 4.4e-10 and *r* = -0.19, *p* = 1.9e-08, respectively). Linear regression analysis of HHQ abundance and individual subjects’ final CLAD stage also showed a significant increase as allograft dysfunction worsened (Pearson *r* = 0.14, *p* = 0.0035) (Figure 3a).

**Figure 3.**
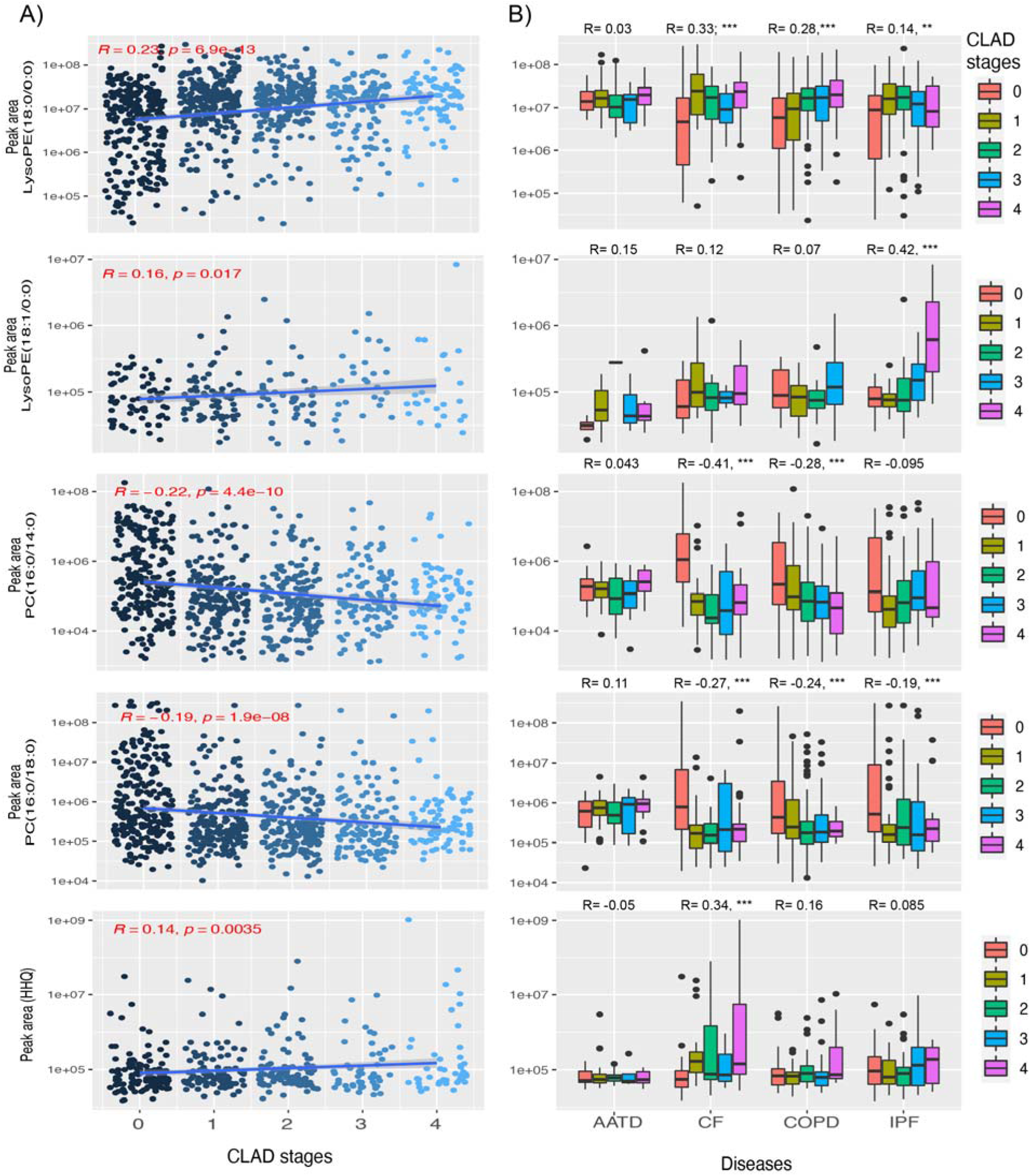
a) Scatter plots and linear regression analysis of feature abundances of target molecules lysoPE (18:0/0:0), lysoPE (18:1/0:0), PC (16:0/14:0), PC(16:0/18:0), and HHQ for all diseases against the final BOS-grade. Statistics of the linear regression are shown on the plots. b) Box plots of feature abundances of each molecule with final CLAD stages separated by individual diseases (AATD, CF, COPD, IPF). Pearson correlation test (R) is displayed, and *** = p values ≤ 0.001.

Finally, we parsed the molecular dynamics of these molecules based on the underlying disease and identified unique trends within each condition. LysoPE (18:0/0:0) significantly increased with disease progression in CF (Pearson *r* = 0.33, p ≤ 0.001), followed by COPD (*r* = 0.28, *p* ≤ 0.001) and IPF (*r* = 0.14 *p* ≤ 0.001), but not in AATD. LysoPE (18:1/0:0) abundance was positively correlated with IPF only (Pearson *r* = 0.42, *p* ≤ 0.001). The feature abundance of PC (16:0/14:0) significantly decreased as disease severity worsened only in CF and COPD (Pearson *r* = -0.41, *p* ≤ 0.001 and *r* = -0.28 *p* ≤ 0.001, respectively), whereas PC (16:0/18:0) significantly decreased in CF, COPD and IPF (Pearson *r* = -0.27, *p* ≤ 0.001; *r* = -0.24, *p* ≤ 0.001; and *r* = -0.19 *p* ≤ 0.001, respectively). Neither of these molecules varied with CLAD stages in AATD. In the case of HHQ, its molecular abundance significantly increased with disease progression in CF (Pearson *r* = 0.34, *p* ≤ 0.001) (Figure 3b) but not the others, indicating that the trend seen in the complete dataset was driven by the CF samples.

### *Pseudomonas aeruginosa* molecular signatures in subjects with CF after LTX

The association of HHQ with CLAD severity led to further analysis of the diversity of *P. aeruginosa* metabolites and their relationship with clinical outcomes, particularly in those subjects with CF. Molecular networking allowed us to identify additional molecular signatures from the pathogen, including 2-heptyl-4-quinolone-N-oxide (HQNO; *m*/*z* 260.1645 [M+H]^+^, C_16_H_22_NO_2_ ; 10.67 ppm), 2-nonyl-4-quinolone (HNQ; *m*/*z* 270.1852 [M+H]^+^, C_17_H_24_NO; 7.45 ppm), pyocyanin (*m*/*z* 211.0866 [M+H]^+^, C_13_H_11_N_2_O ; 5.64 ppm) and pyochelin (*m*/*z* 325.0675 [M+H]^+^, C_14_H_16_N_2_O_3_S_2_; 10.46 ppm). Molecular network node mapping based on disease showed that the entire molecular family of quinolones and pyochelin were enriched in subjects with underlying CF (pie charts within nodes) (Figure 4a). We were therefore interested in determining when these molecules were detected in the longitudinal BALF samples of each subject. We found that ∼60% of samples (n = 46) from subjects with CF were positive for at least one *P. aeruginosa* molecule in the first BALF sample collected (Figure 4b), and the abundances were particularly high at this first-time point (Figure 4c). This proportion of positive samples stayed relatively stable, with a slight decrease in successive BALFs collected (Figure 4b). Some subjects showed acquisition of *P. aeruginosa* molecules as time since transplant progressed, whereas most initially had high abundances and a subsequent decrease (Figure 4c). However, a Pearson correlation between the abundance of these molecules and the time since LTX was not significant (R = 0.074; p = 0.18). The abundance of these molecules did increase (R = 0.34; p = 0.0046) with the relative abundance of *Pseudomonas* sp. (Figure 4d,e).

**Figure 4.**
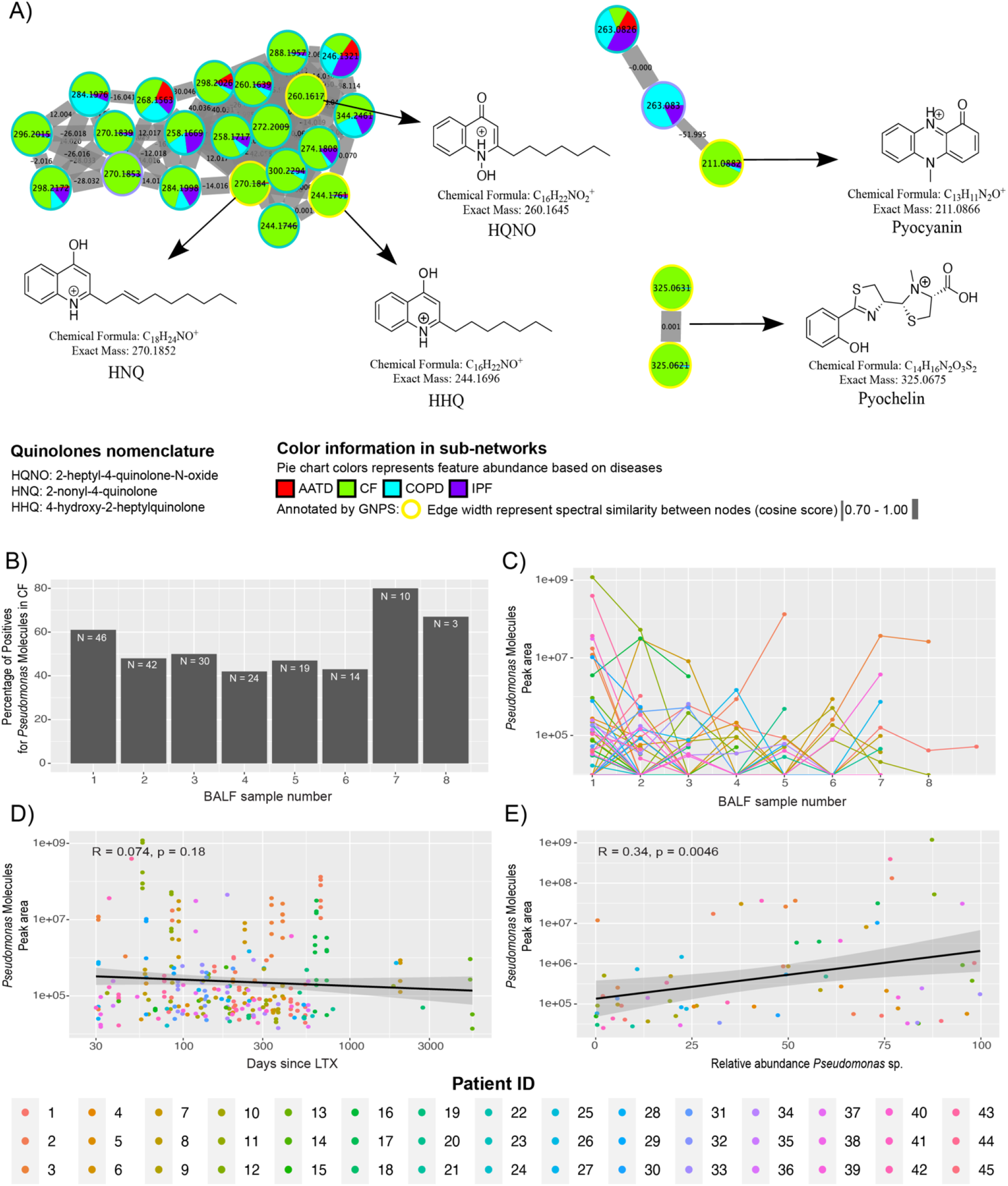
*Pseudomonas*-derived molecules in subjects that underwent lung transplant due to CF disease. A) Molecular networks of microbial molecules produced by *Pseudomonas* sp. that were identified by GNPS library searching. Each node represents a unique MS/MS spectrum (putative molecule), and connections between nodes are determined by spectral similarity (cosine score) from MS/MS alignment. Pie charts represent total feature abundance colored by the underlying disease. B) Percentage of samples that displayed *Pseudomonas*-like molecules in subjects with CF across longitudinal BALF sampling time points and C) dynamics among subjects over time. D) Linear correlation plot displaying the abundance of selected *Pseudomonas* molecules post-LTX and E) its relation to *Pseudomonas spp*. relative abundance in samples.

## Discussion

In this study, we applied untargeted metabolomics to 960 BALF samples from patients who had undergone LTX for chronic lung disease. We had a particular interest in metabolite variation based on the underlying disease and the “gold standard” measure of disease severity (i.e., CLAD stage). The post-LTX metabolome data differed based on the underlying disease, with the unique profile found in AATD. Other chronic lung diseases (CF, COPD, PF) were significantly different from one another but more difficult to distinguish overall. The uniqueness of AATD was driven by differences in aromatic amino acids (phenylalanine), an overall lack of shared molecules with the other diseases, and a lower abundance of metabolite features overall. We note that all four underlying conditions have unique etiologies, and the findings here indicate that, somewhat unexpectedly, the chemical environment of the lung allograft reflects the initial disease of the recipient, particularly in the case of AATD. The high abundance of phenylalanine in allografts from subjects with AATD may reflect increased proteolysis, which is a hallmark of this disease [23], though further research is needed to confirm this hypothesis.

The overall annotation rate of MS/MS spectra in our dataset was 7.6%, which is not uncommon in untargeted metabolomics studies [34]. Low levels of metabolite identification are a known challenge in metabolomics and can limit the ability to infer mechanistic associations with disease severity, although annotation rates in untargeted metabolomics experiments are increasing as new search algorithms and databases become available [22,35–37]. Nevertheless, there is considerable power in using comprehensive molecular data from untargeted metabolomics experiments to identify biological trends, even when the molecular structures are not known. An important step for this approach, which we performed here, is the removal of pharmaceuticals and xenobiotics, which can be highly abundant and overwhelm underlying biological signals. The molecular networking algorithm applied here greatly increases the ability to ‘clean up’ the metabolome by enabling the identification and visualization of pharmaceuticals and their chemical relatives.

Bronchoalveolar lavage fluids have been used recently for describing molecular changes in lipids and metabolites after LTX [16]; however, the association of these compounds with allograft dysfunction has not been addressed. One of the strongest signatures in our metabolomic data was the association of the metabolome with the final CLAD stage of each subject, and this was particularly strong for those with CF. In this case, many of the molecular features driving the association had annotations in the GNPS libraries, and they were primarily phospholipids and bacterial metabolites. A closer analysis of these molecules identified a transition in lipid species associated with an increased final CLAD stage. Subjects whose final CLAD severity was lower had more abundant phosphocholine lipids in their longitudinal BALF metabolome, while those with more severe CLAD outcomes had a higher abundance of phosphoethanolamine lipids. This phospholipid lipid transition may indicate a disruption in airway surface liquid (ASL) in the transplanted lung as the disease progresses or alterations in lipid metabolism. ASL is mainly composed of phospholipids (90%) and proteins (10%), which are products of surface and submucosal gland epithelia and resident phagocytic cells [38]. Approximately 70–80% of ASL lipids are dipalmitoyl phosphatidylcholine (DPPC) [39,40] and PE is a major phospholipid in lung surfactant. The increase in PE abundance associated with CLAD severity found here may be due to the influx of neutrophils or other inflammatory cells containing this lipid. Another potential source is allograft-colonizing microbiota, as PE lipids are known to be a component of bacterial membranes [41,42].

Detection of quorum-sensing metabolites from *P. aeruginosa* in BALF and the association of HHQ with final CLAD severity implicates the bacterium and its metabolism in disease progression. These molecules were particularly prevalent in subjects with underlying CF, where this bacterium is a known opportunistic pathogen responsible for chronic airway infection [43–45] [43]. *P. aeruginosa* regulates the production of its virulence factors through its quorum sensing system, which plays an important role in CF pathogenesis. Multiple quinolones from *P. aeruginosa* such as HQNO, HNQ, and PQS (Pseudomonas quinolone signal), were detected in BALF, as were other two other small molecule virulence factors pyocyanin and pyochelin. Quorum sensing is mediated through N-acyl-homoserine lactones (AHLs) and alkyl quinolones (AQ) [46], the latter of which were detected in this study. The correlation between the abundance of these molecules and the final CLAD stage indicates they could potentially be explored as biomarkers of bacterial infection in CF patients post-LTX and as indicators of CLAD progression. Furthermore, they are relatively easy to detect with LC-MS/MS rapidly after sample extraction. Finally, we note that many of these *P. aeruginosa* small molecules were detected in the first BALF sample after LTX. This observation supports the hypothesis that the bacterium readily re-colonizes the respiratory tract post-LTX from its reservoir in the upper airways (i.e., paranasal sinuses) [44]. Though data from this study cannot directly assess this infection reservoir, the detection of quinolones in BALF so early after LTX is an important finding for understanding the pathogenesis of CLAD.

## Conclusions

The BALF metabolome after lung transplant revealed differences based on underlying lung disease type and association with final CLAD severity. An important metabolic trend was a shift in the relative abundance of phosphocholine and phosphoethanolamine lipids that were predictive of a subject’s final CLAD stages with CF. These findings indicate potential predictive value for the lipid profile as an indicator of disrupted airway surface liquid and impending CLAD severity. Importantly, our LC-MS/MS approach readily detected virulence metabolites from the bacterial pathogen *P. aeruginosa*, especially in CF samples, which were associated with poor CLAD outcomes. This study provides a picture of the molecular transitions within the transplanted lung from the host and bacterial pathogens that may help understand the progression of allograft dysfunction and merits further study with more targeted approaches for the molecules identified.

## Data Availability

All data produced in the present work are contained in the manuscript.

## Acknowledgment

We thank Anthony Schilmiller for his constant support during sample processing in the mass spectrometry and metabolomics core laboratory at Michigan State University. The CF Foundation grant HUNTER18ABO and the National Institute of Allergy and Infectious Diseases (R01AI145925) provided funding.

## Author contributions

R. Hunter and R. Quinn conceptualized the study. K. Mahan performed BALF/CLAD stages tabulation. C. Martin, T. Wiggen, and R. Quinn performed data acquisition, analysis, and interpretation. C. Martin, R. Quinn, and R. Hunter wrote the original and final versions of the manuscript. K. Mahan and M. Hertz edited the manuscript. All authors read and approved the final manuscript.

