## Supplementary Information for "Bronchoalveolar lavage metabolome dynamics reflect underlying disease and chronic lung allograft dysfunction"

**Supplementary methods**

**Organic extraction and LC-MS/MS analysis**

Individually, BALF samples (500 uL) from 194 patients with CLAD associated-diseases (supplementary table 2) were suspended in methanol (500 uL) LC-MS grade into a microcentrifuge tube (1.50 mL). These tubes were vortexed for 30 seconds and centrifuged at 3000 g for 10 minutes. Then, tubes were dried by means of a speed vac for 3 - 4 hours or until obtaining a dried pellet. Pellets into microcentrifuge tubes were reconstituted in methanol LC-MS grade (120 uL). From those, 100 uL were placed on 96 well plates and 10 uL were used for pool quality controls [1] (supplementary figure 1).

The resulting plates were analyzed on a Thermo™ QExactive™ mass spectrometer coupled to a Vanquish Ultra-High-Performance Liquid Chromatography (UHPLC) system. The mobile phase was 0.1% formic acid in Milli-Q water (channel A) and acetonitrile (channel B). The stationary phase was a reverse-phase column Waters® Acquity® (Wood Dale, IL, USA) UPLC BEH C-18 column, 2.1 mm × 100 mm. The chromatographic runs were 12 min-long with linear gradients as follows: 0–1 min 2% B, 1–8 min 2–100% B. This 100% B solution was then held for 2 min followed by a switch to 2% B for the remaining 2 min. The injection volume was 10 µL, the flow rate 0.40 mL/min, and the column temperature 60°C. Full MS1 survey scans and MS2 mass spectra for five precursor ions per survey scan were collected using electrospray ionization in positive mode with a scan range set from m/z 100 to 1500 for the full MS mode (minutes 1–10 of run) [2]. Raw files (.raw) were converted to .mzXML format for processing and analyzing with MZmine 2.53 software (supplementary table 3) and GNPS [3,4].

### MS/MS molecular networking

Feature-based molecular networking was generated using the Global Natural Products Social Molecular Networking online platform (GNPS). The precursor ion mass tolerance was set to 0.02 Da and the MS/MS fragment ion tolerance to 0.02 Da [3,5]. A molecular network was then created where edges were filtered to have a cosine score above 0.70 and more than 4 matched peaks. Further, edges between two nodes were kept in the network if and only if each of the nodes appeared in each other's respective top 10 most similar nodes. Finally, the maximum size of a molecular family was set to 100, and the lowest-scoring edges were removed from molecular families until the molecular family size was below this threshold. The spectra in the network were then searched against GNPS spectral libraries [3,6]. The library spectra were filtered in the same manner as the input data. All matches kept between network spectra and library spectra were required to have a score above 0.70 and at least 4 matched peaks. The DEREPLICATOR was used to annotate MS/MS spectra [7]. The resulting molecular network was imported to cytoscape 3.8.0 (https://cytoscape.org/) and analyzed using default algorithms and other visual considerations [8,9].

### Statistical analysis

Data curation was done considering both, the feature quantification table generated by mzMine2 and the molecular annotations obtained from GNPS. First, we removed blank extraction signals after analyzing their feature intensity in the raw data (blank extraction samples) and then set a filter threshold in the molecular network. This filter was set for blank extraction controls from $5.0e^5$ to $1.5e^8$. All selected nodes were removed from the feature quantification table. Additionally, we did a detailed removal of those nodes annotated by GNPS as drugs and also those non-annotated nodes which were directly related to drugs in the molecular network. After blanks, drugs, and drugs-related data removal, we generated a bucket table that basically includes all biological signals generated from BALF samples. Thus, the variation in the metabolomic data associated with disease type and CLAD stage was assessed using beta-diversity metrics with

PERMANOVA testing and machine-learning-based random forest (RF) classification and regression analysis. This dual analysis approach was done on the entire dataset and each disease individually. A Bray-Curtis dissimilarity matrix was calculated on the entire metabolome containing MS/MS feature intensities for all samples and used to generate principal coordinate analysis (PCoA) plots. These plots were created through the in-house tool ClusterApp and visualized in EMPeror (10). PERMANOVA tests were performed for diseases and CLAD stages (as a categorical variable) with patient-source as an interacting factor to account for the variation in the number of samples per patient. Random forest (RF) analysis with 1,000 decision trees was used to classify the metabolome based on disease type (RF classification) and CLAD stage (RF regression). Out-of-bag (OOB) errors were reported for classification and the percent of variance explained by the CLAD stage for the regressions to assess the influence of these CLAD measures on the metabolomic data. RF analysis was also run on a reduced bucket table containing only those MS/MS features identified as known by GNPS library searching. Variable importance plots from both RF approaches were used to identify metabolites driving the variation observed. These random forests were run on the combined dataset and for each disease separately. Pearson correlations were used to determine the relationship between the abundance of metabolites of interest and CLAD stage (linearized variable). Additionally, we evaluated the longitudinal changes in the BALF metabolome and disease severity (CLAD stage) through time in a sub-group of patients (n = 78) who had at least 6 samples. Only those 6 samples collected chronologically were analyzed in this manner to normalize across the subjects. The R packages vegan and ggplot2 were used for these analyses (11-13).

**Supplementary figures**

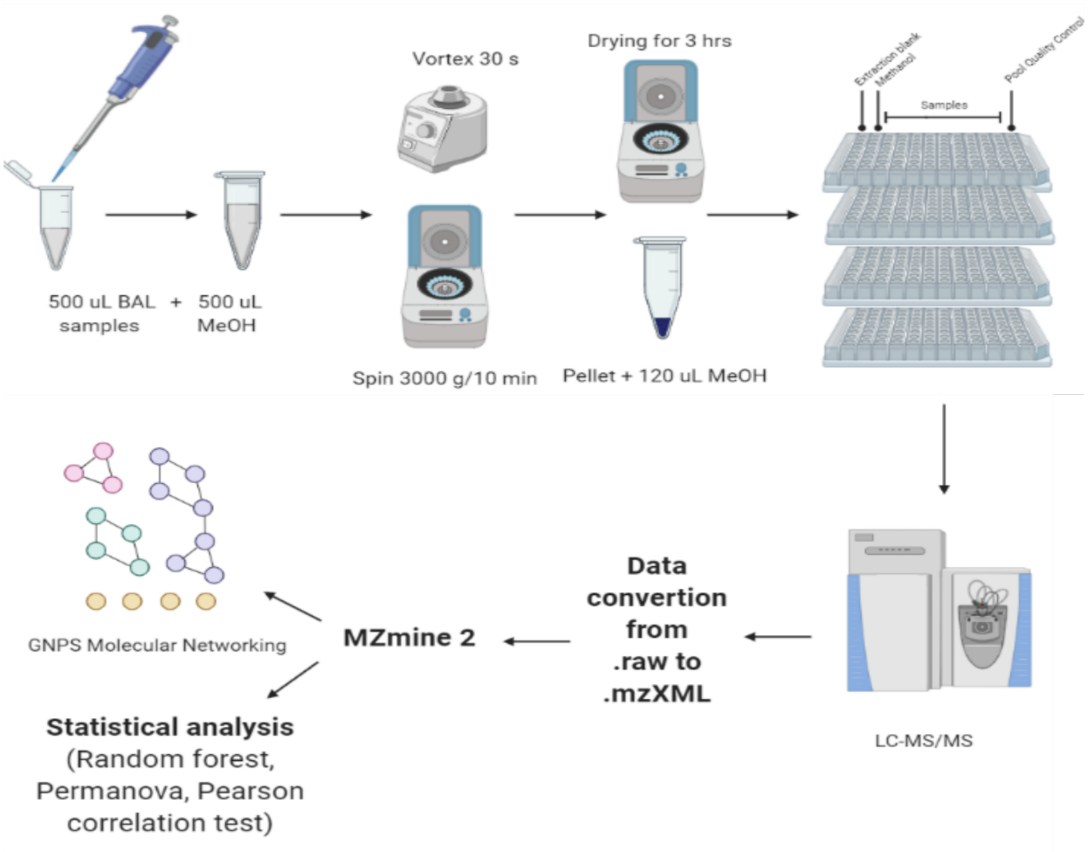


**Supplementary figure 1** Sample processing workflow for metabolomic data acquisition in bronchoalveolar lavage samples.

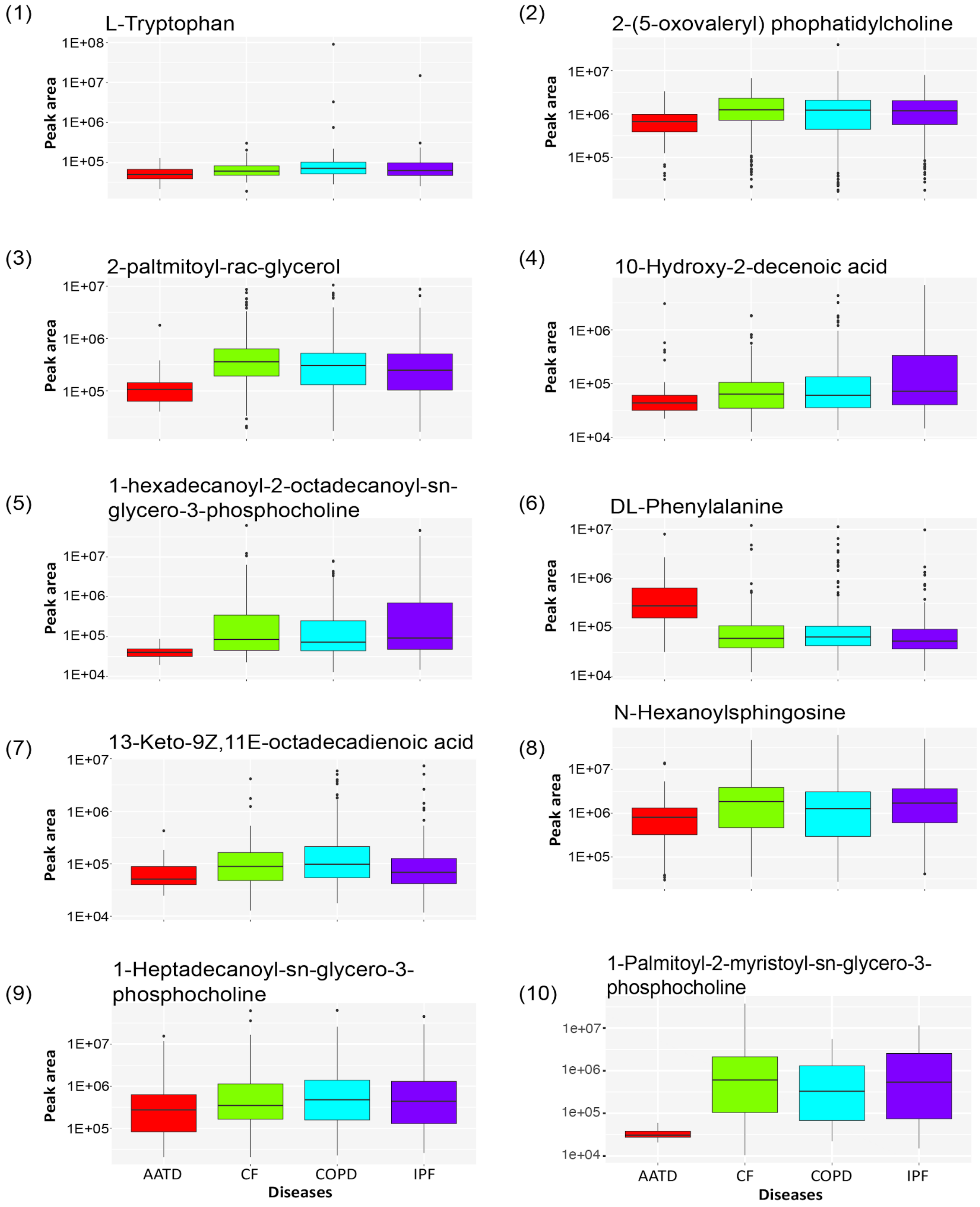


**Supplementary figure 2** Box plots displaying the feature abundance across pre-transplant diagnosed diseases in BALF samples. 1-10 represents the top ten ranked molecules in the variable importance plot generated from supervised regression random forest analysis in AATD and its dynamics across diseases.

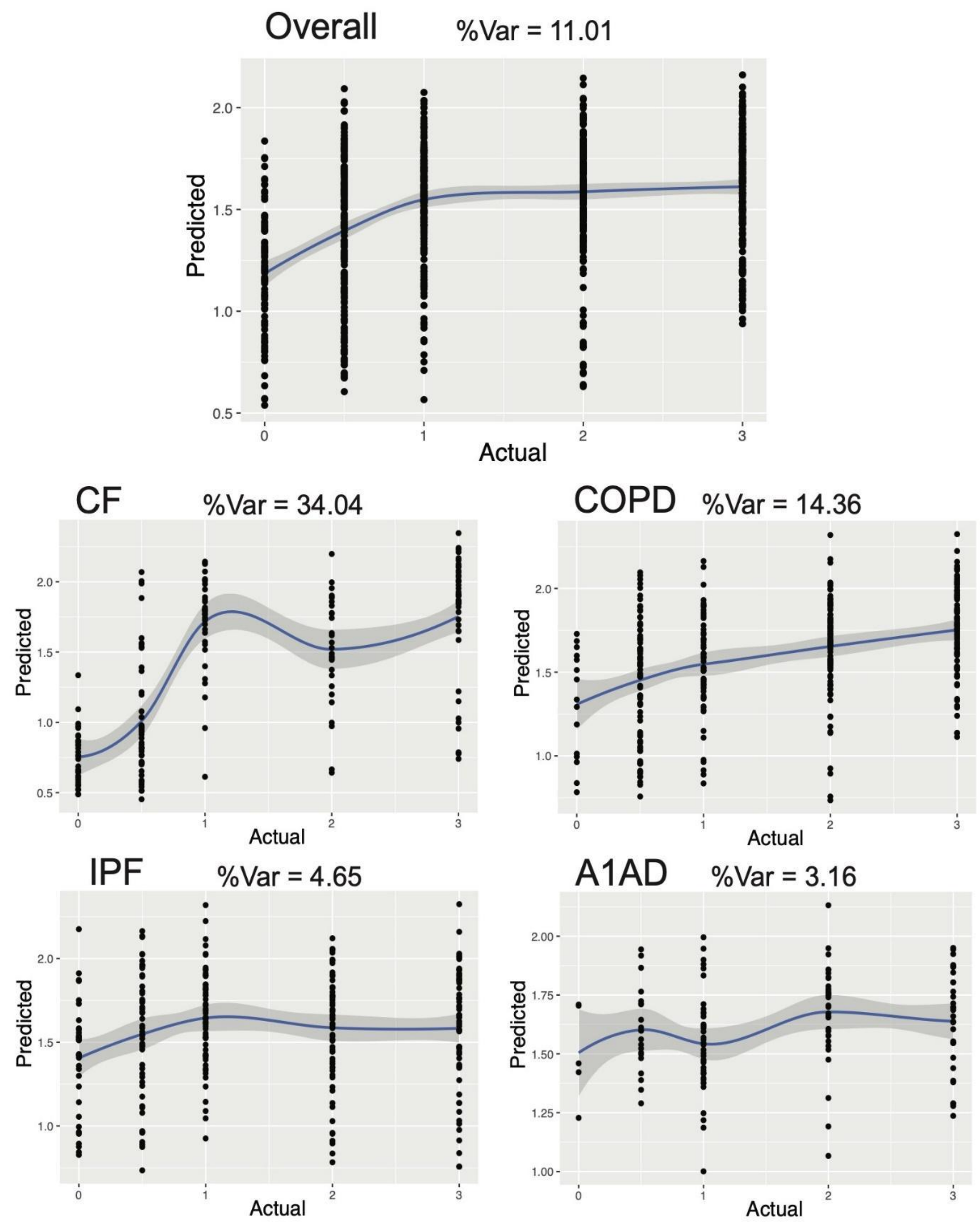


**Supplementary figure 3** Scatterplot of the CLAD stage for each sample and the predicted score from the random forest regression based on the metabolomic data. The plots are for the entire dataset together with all diseases (overall) and the individual diseases separately. The smoothed mean line and its 95% confidence interval are also plotted. The percent of variance explained by the CLAD stage in the RF regression is shown.

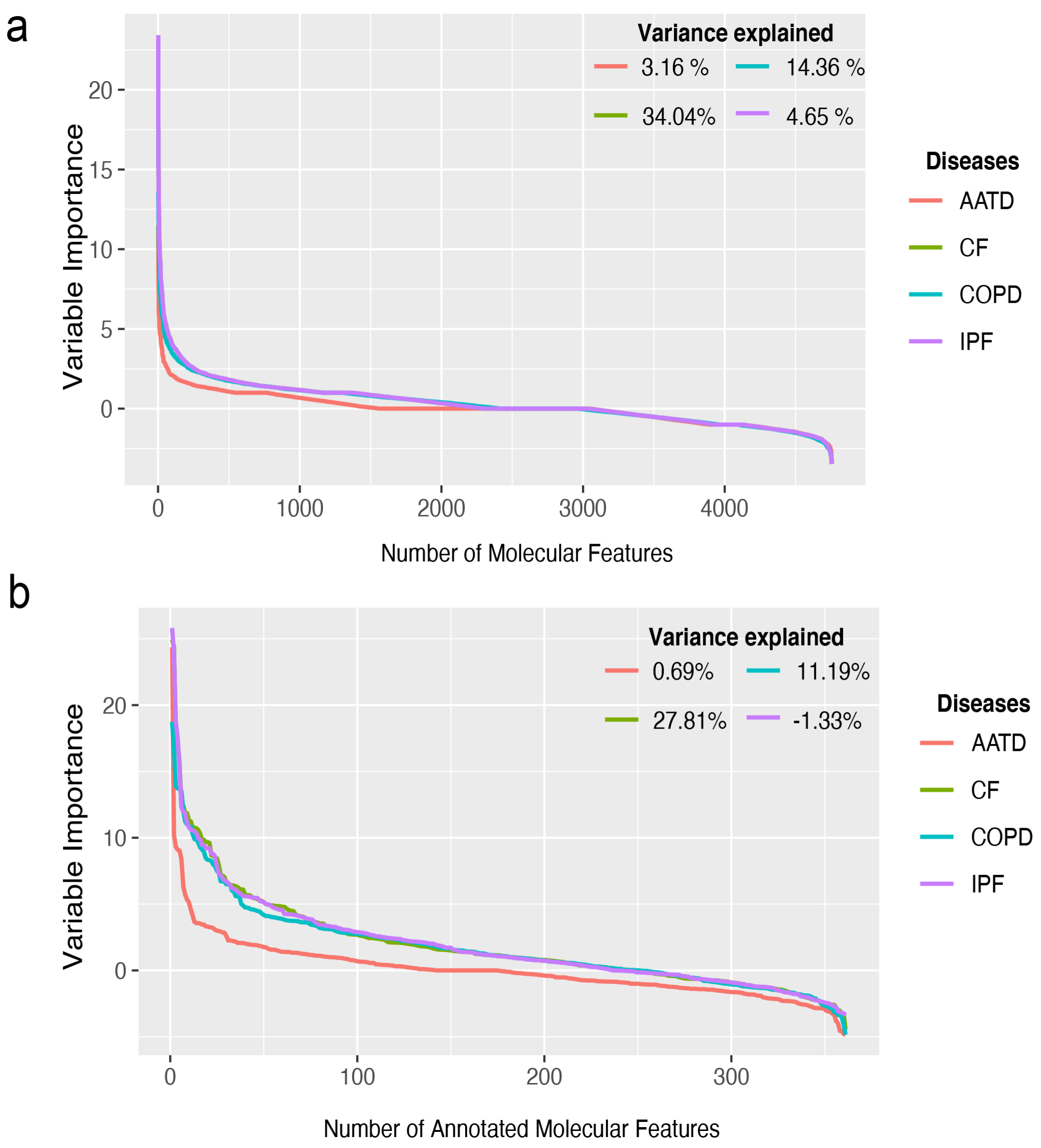


**Supplementary figure 4** Variance importance plots from supervised regression random forest of a) BALF metabolome and b) BALF-annotated metabolome (annotated molecules by GNPS) based on the CLAD stage in the individual diseases AATD, CF, COPD, and IPF. The percentage of variance explained in both molecular features, BALF and BALF-annotated metabolome, indicates a greater effect on the increase of CLAD stage in CF (34.04%, 27.81%) and COPD (14.36%,11.19%) respectively.

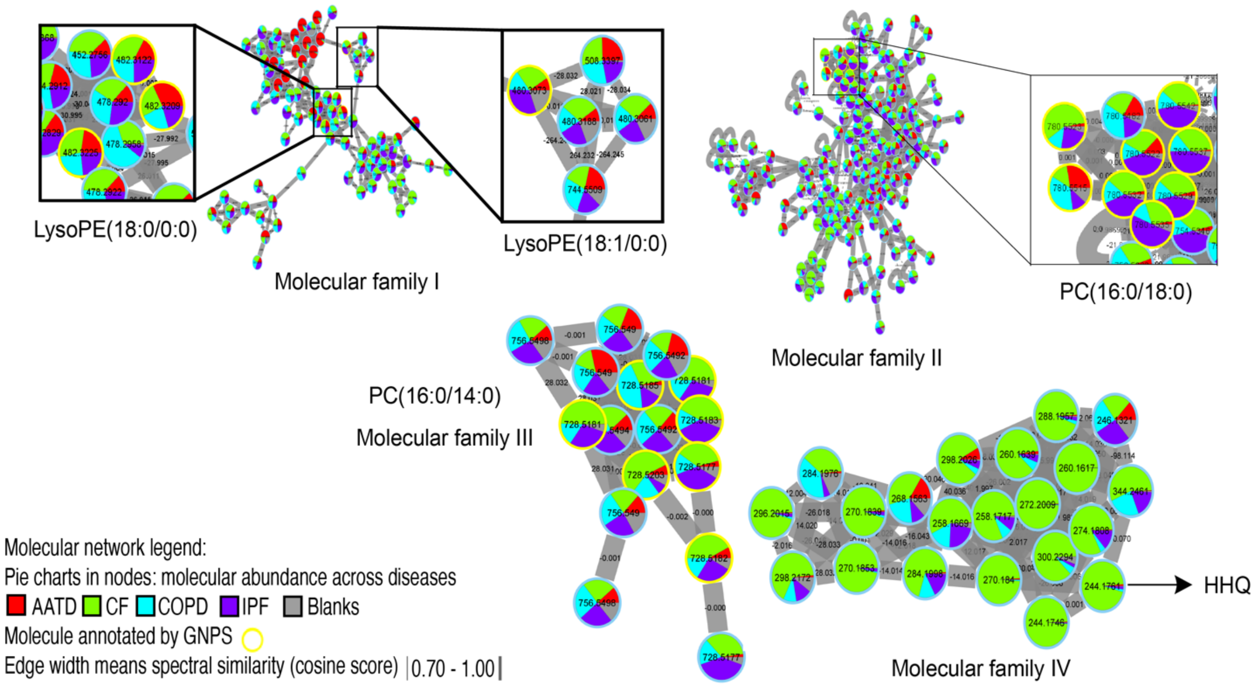


**Supplementary figure 5** Molecular networks associated with changes between CLAD stage and diseases. These networks correspond to molecular families, namely, phosphoethanolamine (I), phosphocholine (II and III), and quinolones (IV). Each node represents a unique MS/MS spectrum (putative molecule), connections between the nodes are scaled to the cosine score and the pie chart denotes the total abundance of that molecule in each of the four diseases. Highlighted nodes (yellow) correspond to those molecules annotated by GNPS (4).

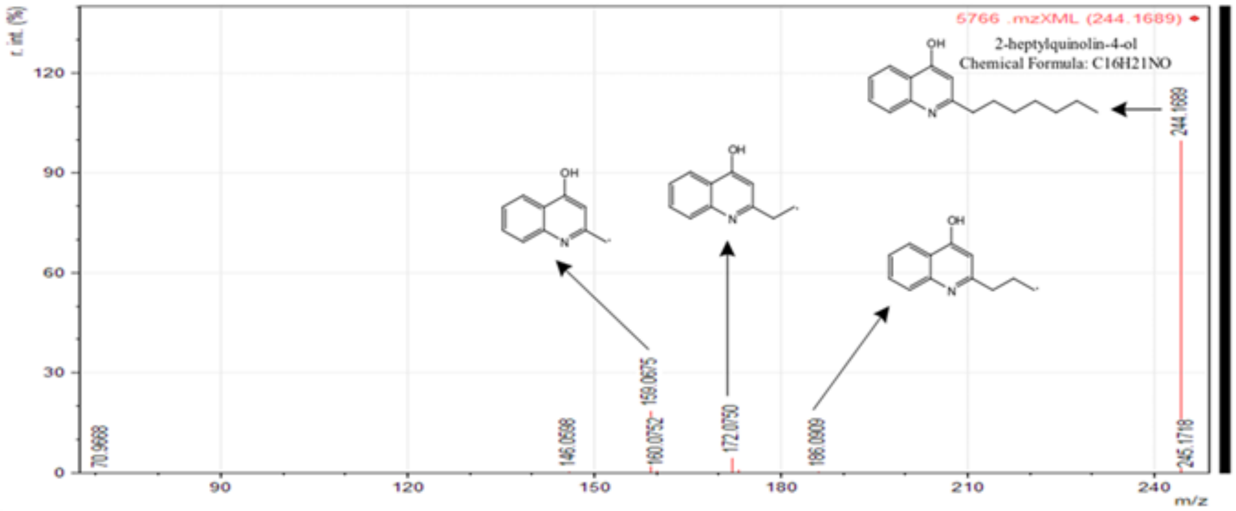


**Supplementary figure 6** Fragmentation pattern displayed by 2-heptyl quinolin-4-ol (HHQ) manually annotated from raw data.

**Supplementary tables**

| Sample No. | Subject ID | No. Samples | Pre-Diagnosed Diseases | Sex | Last CLAD stage | Last Avg $FEV_1$ |
|---|---|---|---|---|---|---|
| 1 | Subject_001 | 5 | AATD | Male | 0 | 85.53 |
| 2 | Subject_002 | 7 | AATD | Female | 1 | 78.52 |
| 3 | Subject_003 | 7 | AATD | Male | 4 | 32.64 |
| 4 | Subject_004 | 4 | AATD | Female | 0 | 87.69 |
| 5 | Subject_005 | 6 | AATD | Male | 1 | 69.09 |
| 6 | Subject_006 | 2 | AATD | Female | 4 | 34.35 |
| 7 | Subject_007 | 8 | AATD | Male | 2 | 54.69 |
| 8 | Subject_008 | 9 | AATD | Male | 3 | 47.77 |
| 9 | Subject_009 | 2 | AATD | Female | 0 | 85.27 |
| 10 | Subject_010 | 2 | AATD | Male | 1 | 79.52 |
| 11 | Subject_011 | 6 | AATD | Male | 1 | 74.12 |
| 12 | Subject_012 | 2 | AATD | Female | 3 | 48.72 |
| 13 | Subject_013 | 6 | AATD | Male | 4 | 18.02 |
| 14 | Subject_014 | 6 | AATD | Male | 0 | 83.16 |
| 15 | Subject_015 | 12 | AATD | Female | 2 | 54.78 |
| 16 | Subject_016 | 4 | AATD | Female | 2 | 59.07 |
| 17 | Subject_017 | 7 | AATD | Male | 2 | 61.03 |
| 18 | Subject_018 | 4 | AATD | Female | 0 | 96.23 |
| 19 | Subject_019 | 6 | AATD | Female | 0 | 81.48 |
| 20 | Subject_020 | 9 | AATD | Male | 1 | 78.03 |
| 21 | Subject_021 | 9 | AATD | Male | 1 | 75.51 |
| 22 | Subject_022 | 7 | AATD | Male | 4 | 28.89 |
| 23 | Subject_023 | 9 | CF | Female | 0 | 96.58 |
| 24 | Subject_024 | 5 | CF | Male | 2 | 51.15 |
| 25 | Subject_025 | 8 | CF | Female | 1 | 79.53 |
| 26 | Subject_026 | 4 | CF | Female | 1 | 67.56 |
| 27 | Subject_027 | 5 | CF | Male | 4 | 22.03 |
| 28 | Subject_028 | 5 | CF | Female | 0 | 85.65 |
| 29 | Subject_029 | 6 | CF | Female | 4 | 25 |
| 30 | Subject_030 | 7 | CF | Male | 1 | 80.41 |
| 31 | Subject_031 | 7 | CF | Male | 1 | 68.19 |
| 32 | Subject_032 | 7 | CF | Female | 2 | 65.43 |
| 33 | Subject_033 | 8 | CF | Male | 2 | 59.78 |
| 34 | Subject_034 | 6 | CF | Female | 4 | 26.32 |
| 35 | Subject_035 | 4 | CF | Female | 0 | 100 |
| 36 | Subject_036 | 2 | CF | Male | 3 | 39.12 |
| 37 | Subject_037 | 5 | CF | Male | 3 | 50.25 |

| | | | | | | |
|---|---|---|---|---|---|---|
| 38 | Subject_038 | 2 | CF | Female | 2 | 57.25 |
| 39 | Subject_039 | 3 | CF | Male | 2 | 60.04 |
| 40 | Subject_040 | 2 | CF | Male | 4 | 20.27 |
| 41 | Subject_041 | 3 | CF | Male | 2 | 62.24 |
| 42 | Subject_042 | 5 | CF | Male | 1 | 77.16 |
| 43 | Subject_043 | 5 | CF | Female | 4 | 25.88 |
| 44 | Subject_044 | 1 | CF | Male | 4 | 26.39 |
| 45 | Subject_045 | 1 | CF | Male | 1 | 71.99 |
| 46 | Subject_046 | 4 | CF | Male | 4 | 16.85 |
| 47 | Subject_047 | 2 | CF | Male | 0 | 85.71 |
| 48 | Subject_048 | 3 | CF | Female | 4 | 33.33 |
| 49 | Subject_049 | 2 | CF | Female | 0 | 87.04 |
| 50 | Subject_050 | 7 | CF | Female | 4 | 28.83 |
| 51 | Subject_051 | 3 | CF | Male | 0 | 98.33 |
| 52 | Subject_052 | 5 | CF | Female | 0 | 89.31 |
| 53 | Subject_053 | 2 | CF | Male | 1 | 71.11 |
| 54 | Subject_054 | 6 | CF | Male | 0 | 83.67 |
| 55 | Subject_055 | 1 | CF | Male | 2 | 60.21 |
| 56 | Subject_056 | 3 | CF | Female | 0 | 98.56 |
| 57 | Subject_057 | 2 | CF | Male | 0 | 86.9 |
| 58 | Subject_058 | 2 | CF | Female | 0 | 83.11 |
| 59 | Subject_059 | 3 | CF | Female | 0 | 90.1 |
| 60 | Subject_060 | 1 | CF | Female | 0 | 94.47 |
| 61 | Subject_061 | 4 | CF | Female | 0 | 90.35 |
| 62 | Subject_062 | 7 | CF | Male | 0 | 93.59 |
| 63 | Subject_063 | 7 | CF | Female | 0 | 89.66 |
| 64 | Subject_064 | 3 | CF | Female | 0 | 89.58 |
| 65 | Subject_065 | 2 | CF | Male | 0 | 92.95 |
| 66 | Subject_066 | 3 | CF | Female | 1 | 77.37 |
| 67 | Subject_067 | 1 | CF | Male | 0 | 83.42 |
| 68 | Subject_068 | 2 | CF | Female | 0 | 83.05 |
| 69 | Subject_069 | 2 | COPD | Male | 0 | 91.16 |
| 70 | Subject_070 | 6 | COPD | Female | 0 | 96.45 |
| 71 | Subject_071 | 3 | COPD | Male | 0 | 100 |
| 72 | Subject_072 | 9 | COPD | Female | 1 | 80.13 |
| 73 | Subject_073 | 7 | COPD | Female | 1 | 79.59 |
| 74 | Subject_074 | 6 | COPD | Female | 1 | 80.25 |
| 75 | Subject_075 | 2 | COPD | Female | 1 | 75.9 |
| 76 | Subject_076 | 2 | COPD | Female | 1 | 70.63 |
| 77 | Subject_077 | 5 | COPD | Male | 1 | 77.78 |

| | | | | | | |
|---|---|---|---|---|---|---|
| 78 | Subject_078 | 3 | COPD | Female | 1 | 74.82 |
| 79 | Subject_079 | 3 | COPD | Female | 1 | 74.29 |
| 80 | Subject_080 | 3 | COPD | Female | 1 | 69.05 |
| 81 | Subject_081 | 6 | COPD | Male | 1 | 69.52 |
| 82 | Subject_082 | 6 | COPD | Female | 1 | 76.88 |
| 83 | Subject_083 | 5 | COPD | Female | 1 | 67.35 |
| 84 | Subject_084 | 4 | COPD | Male | 1 | 72.12 |
| 85 | Subject_085 | 5 | COPD | Male | 1 | 79.06 |
| 86 | Subject_086 | 8 | COPD | Male | 1 | 68.02 |
| 87 | Subject_087 | 6 | COPD | Female | 2 | 56.03 |
| 88 | Subject_088 | 6 | COPD | Male | 2 | 65.24 |
| 89 | Subject_089 | 5 | COPD | Male | 2 | 51.85 |
| 90 | Subject_090 | 8 | COPD | Male | 2 | 53.8 |
| 91 | Subject_091 | 6 | COPD | Male | 2 | 56.16 |
| 92 | Subject_092 | 2 | COPD | Female | 2 | 52.28 |
| 93 | Subject_093 | 2 | COPD | Male | 2 | 56.04 |
| 94 | Subject_094 | 7 | COPD | Female | 2 | 63.06 |
| 95 | Subject_095 | 6 | COPD | Male | 2 | 53.51 |
| 96 | Subject_096 | 4 | COPD | Female | 2 | 52.17 |
| 97 | Subject_097 | 5 | COPD | Female | 2 | 58.06 |
| 98 | Subject_098 | 6 | COPD | Male | 2 | 55.71 |
| 99 | Subject_099 | 3 | COPD | Female | 2 | 58.18 |
| 100 | Subject_100 | 6 | COPD | Male | 2 | 58.11 |
| 101 | Subject_101 | 6 | COPD | Female | 2 | 61.57 |
| 102 | Subject_102 | 4 | COPD | Male | 2 | 57.85 |
| 103 | Subject_103 | 5 | COPD | Female | 2 | 62.25 |
| 104 | Subject_104 | 2 | COPD | Female | 2 | 59.21 |
| 105 | Subject_105 | 4 | COPD | Female | 2 | 58.4 |
| 106 | Subject_106 | 3 | COPD | Female | 2 | 60.79 |
| 107 | Subject_107 | 5 | COPD | Female | 4 | 31.28 |
| 108 | Subject_108 | 6 | COPD | Female | 3 | 37.07 |
| 109 | Subject_109 | 6 | COPD | Male | 3 | 35.97 |
| 110 | Subject_110 | 4 | COPD | Male | 3 | 46.58 |
| 111 | Subject_111 | 4 | COPD | Female | 3 | 44.44 |
| 112 | Subject_112 | 7 | COPD | Male | 3 | 36.14 |
| 113 | Subject_113 | 3 | COPD | Male | 3 | 40.3 |
| 114 | Subject_114 | 5 | COPD | Female | 4 | 31.5 |
| 115 | Subject_115 | 7 | COPD | Female | 3 | 37.93 |
| 116 | Subject_116 | 2 | COPD | Male | 3 | 44.9 |
| 117 | Subject_117 | 2 | COPD | Male | 4 | 22.71 |

| | | | | | | |
|---|---|---|---|---|---|---|
| 118 | Subject_118 | 5 | COPD | Female | 3 | 38.16 |
| 119 | Subject_119 | 3 | COPD | Female | 3 | 38.21 |
| 120 | Subject_120 | 8 | COPD | Male | 3 | 43.69 |
| 121 | Subject_121 | 2 | COPD | Female | 4 | 27.27 |
| 122 | Subject_122 | 5 | COPD | Female | 3 | 47.4 |
| 123 | Subject_123 | 6 | COPD | Female | 3 | 41.1 |
| 124 | Subject_124 | 4 | COPD | Male | 3 | 46.6 |
| 125 | Subject_125 | 5 | COPD | Female | 3 | 44.44 |
| 126 | Subject_126 | 3 | COPD | Male | 3 | 38.29 |
| 127 | Subject_127 | 4 | COPD | Female | 3 | 40.15 |
| 128 | Subject_128 | 4 | COPD | Male | 0 | 87.16 |
| 129 | Subject_129 | 6 | COPD | Female | 0 | 82.74 |
| 130 | Subject_130 | 3 | COPD | Male | 0 | 80.56 |
| 131 | Subject_131 | 7 | COPD | Female | 0 | 85.71 |
| 132 | Subject_132 | 5 | COPD | Male | 0 | 84.32 |
| 133 | Subject_133 | 6 | COPD | Male | 0 | 90.34 |
| 134 | Subject_134 | 2 | COPD | Female | 0 | 80.82 |
| 135 | Subject_135 | 6 | COPD | Female | 0 | 90.85 |
| 136 | Subject_136 | 4 | COPD | Female | 0 | 94.03 |
| 137 | Subject_137 | 2 | COPD | Male | 0 | 100 |
| 138 | Subject_138 | 2 | COPD | Female | 0 | 88.73 |
| 139 | Subject_139 | 2 | COPD | Male | 0 | 90.74 |
| 140 | Subject_140 | 6 | COPD | Male | 0 | 84.65 |
| 141 | Subject_141 | 4 | COPD | Male | 0 | 91.8 |
| 142 | Subject_142 | 7 | COPD | Male | 0 | 92.83 |
| 143 | Subject_143 | 4 | COPD | Male | 0 | 86.83 |
| 144 | Subject_144 | 6 | COPD | Female | 0 | 85.27 |
| 145 | Subject_145 | 3 | COPD | Female | 0 | 89.58 |
| 146 | Subject_146 | 2 | COPD | Female | 0 | 82.47 |
| 147 | Subject_147 | 4 | COPD | Male | 0 | 91.8 |
| 148 | Subject_148 | 3 | IPF | Male | 2 | 62.61 |
| 149 | Subject_149 | 10 | IPF | Male | 3 | 38.55 |
| 150 | Subject_150 | 8 | IPF | Female | 1 | 75.35 |
| 151 | Subject_151 | 8 | IPF | Male | 2 | 63.23 |
| 152 | Subject_152 | 5 | IPF | Male | 0 | 94.94 |
| 153 | Subject_153 | 8 | IPF | Male | 2 | 51.1 |
| 154 | Subject_154 | 8 | IPF | Male | 0 | 88.94 |
| 155 | Subject_155 | 3 | IPF | Male | 2 | 65.06 |
| 156 | Subject_156 | 7 | IPF | Female | 1 | 68.16 |
| 157 | Subject_157 | 7 | IPF | Female | 1 | 68.6 |

| | | | | | | |
|---|---|---|---|---|---|---|
| 158 | Subject_158 | 6 | IPF | Male | 3 | 41.35 |
| 159 | Subject_159 | 7 | IPF | Male | 3 | 41.83 |
| 160 | Subject_160 | 3 | IPF | Male | 1 | 72.47 |
| 161 | Subject_161 | 6 | IPF | Female | 1 | 67 |
| 162 | Subject_162 | 5 | IPF | Male | 2 | 53.24 |
| 163 | Subject_163 | 7 | IPF | Male | 2 | 64.33 |
| 164 | Subject_164 | 5 | IPF | Female | 0 | 84.77 |
| 165 | Subject_165 | 3 | IPF | Female | 2 | 59.49 |
| 166 | Subject_166 | 6 | IPF | Male | 0 | 98.72 |
| 167 | Subject_167 | 5 | IPF | Female | 1 | 68.61 |
| 168 | Subject_168 | 7 | IPF | Male | 4 | 26.37 |
| 169 | Subject_169 | 7 | IPF | Male | 0 | 83.12 |
| 170 | Subject_170 | 8 | IPF | Female | 0 | 86.52 |
| 171 | Subject_171 | 6 | IPF | Male | 1 | 80.21 |
| 172 | Subject_172 | 6 | IPF | Male | 2 | 62.9 |
| 173 | Subject_173 | 4 | IPF | Female | 0 | 85.87 |
| 174 | Subject_174 | 7 | IPF | Male | 1 | 78.17 |
| 175 | Subject_175 | 7 | IPF | Male | 1 | 68.03 |
| 176 | Subject_176 | 7 | IPF | Male | 1 | 68.56 |
| 177 | Subject_177 | 6 | IPF | Male | 2 | 56.74 |
| 178 | Subject_178 | 9 | IPF | Male | 2 | 50.68 |
| 179 | Subject_179 | 8 | IPF | Male | 2 | 56.71 |
| 180 | Subject_180 | 3 | IPF | Male | 3 | 36.05 |
| 181 | Subject_181 | 3 | IPF | Male | 1 | 78.87 |
| 182 | Subject_182 | 9 | IPF | Female | 3 | 36.13 |
| 183 | Subject_183 | 5 | IPF | Male | 0 | 82.35 |
| 184 | Subject_184 | 5 | IPF | Female | 0 | 96.41 |
| 185 | Subject_185 | 6 | IPF | Male | 0 | 97.7 |
| 186 | Subject_186 | 5 | IPF | Female | 2 | 63.8 |
| 187 | Subject_187 | 9 | IPF | Female | 0 | 83.42 |
| 188 | Subject_188 | 6 | IPF | Male | 0 | 91.39 |
| 189 | Subject_189 | 7 | IPF | Male | 3 | 43.43 |
| 190 | Subject_190 | 6 | IPF | Female | 1 | 79.08 |
| 191 | Subject_191 | 5 | IPF | Male | 4 | 27.44 |
| 192 | Subject_192 | 6 | IPF | Male | 0 | 82.78 |
| 193 | Subject_193 | 3 | IPF | Male | 0 | 93.14 |
| 194 | Subject_194 | 3 | IPF | Male | 0 | 92.81 |

126

**Supplementary table 1 –** Study subjects, CLAD associated-diseases and disease progression based on CLAD stages. CLAD stages are from 0-4; where 0 represent the forced expiratory volume in 1 second ($FEV_1$) > 80%, 1 represent $FEV_1$ = 65-80%, 2 means $FEV_1$ = 50-65%, 3 correspond to $FEV_1$ = 35-50% and 4 denote FEV1 ≤ 35% [10]. Additional subject information for research is also displayed.

| Procedure | Description | Parameter |
|---|---|---|
| Mass detection | MS1 | 1.0E5 |
| | MS2 | 1.0E3 |
| | Mass detector | Centroid |
| ADAP chromatograms | Minimum group size | 5 |
| | Group intensity | 3.0E6 |
| | Minimum highest intensity | 3.0E6 |
| | *m/z* tolerance | 0.02 Da or 10 ppm |
| Chromatogram deconvolution | Algorithm | Baseline cut-off |
| | Minimum peak height | 3.0E6 |
| | Peak duration range (min) | 0.10-1.0 |
| | Baseline level | 1.0E6 |
| Isotopic peak grouper | *m/z* tolerance | 0.02 Da or 10 ppm |
| | Retention time tolerance (min) | 0.10 |
| | Maximum charge | 2 |
| Join alignment | *m/z* tolerance | 0.02 Da or 10 ppm |
| | Weight for *m/z* | 75 |

| | | |
|---|---|---|
| | Weight for retention time | 25 |
| | Retention time tolerance (absolute: min) | 0.10 |
| Duplicate peak filter | *m/z* tolerance | 0.02 Da or 10 ppm |
| | Retention time tolerance (min) | 0.10 |
| Gap-filled | Intensity tolerance | 0.05 |
| | *m/z* tolerance | 0.02 Da or 10 ppm |
| | Retention time tolerance (min) | 0.10 |
| | Require same charge state | True |
| Peak list row filter | Minimum peak in row | 5 |
| | Minimum peak isotope | 1 |
| | Reset number ID | True |
| | Keep only peaks with $MS^2$ scan (GNPS) | True |
| MetaCorrelate | Retention time tolerance (min) | 0.10 |
| | Minimum peak height | 3.0E6 |
| | Noise level | 1.0E6 |

**Supplementary table 2** Parameters applied to mzXML converted files in mzMine2 software for performing feature-based molecular networking.

| | A1AD | CF | COPD | IPF | Class. Error |
|---|---|---|---|---|---|
| AATD | 111 | 0 | 15 | 2 | 13.13% |
| CF | 0 | 53 | 100 | 36 | 72.0% |
| COPD | 3 | 8 | 268 | 73 | 23.9% |
| IPF | 0 | 2 | 152 | 130 | 54.2% |

**Supplementary Table 3** Confusion matrix of RF classification of metabolomic data based on disease source. The overall out-of-bag error was 41.03%.

| **Diseases** | **F-value** | **p-value** |
|---|---|---|
| A1AD | 6.1197 | 0.001 |
| CF | 7.5132 | 0.001 |
| COPD | 2.9641 | 0.005 |
| IPF | 3.3776 | 0.002 |

**Supplementary table 4** PERMANOVA test results of patient-specific signature in the metabolome identified by GNPS among all patients and individual diseases**.**

5. Nothias LF, Petras D, Schmid R, et al. Feature-based Molecular Networking in the GNPS Analysis Environment. doi:10.1101/812404

6. Nothias LF, Petras D, Schmid R, et al. Feature-based Molecular Networking in the GNPS Analysis Environment. doi:10.1101/812404

7. Mohimani H, Gurevich A, Shlemov A, et al. Dereplication of microbial metabolites through database search of mass spectra. *Nat Commun*. 2018;9(1):1-12.

8. Shannon P, Markiel A, Ozier O, et al. Cytoscape: a software environment for integrated models of biomolecular interaction networks. *Genome Res*. 2003;13(11):2498-2504.

9. Martin H C, Ibáñez R, Nothias LF, et al. Viscosin-like lipopeptides from frog skin bacteria inhibit Aspergillus fumigatus and Batrachochytrium dendrobatidis detected by imaging mass spectrometry and molecular networking. *Sci Rep*. 2019;9(1):3019.

10. Verleden GM, Glanville AR, Lease ED, *et al.* Chronic lung allograft dysfunction: Definition, diagnostic criteria, and approaches to treatment- A consensus report from the Pulmonary Council of the ISHLT. *J Heart Lung Transplant* 2019;**38**:493–503.
